# Detection of Danger-Associated Molecular Patterns in Kidney Preservation Fluid as Predictors of Delayed Graft Function after Transplantation

**DOI:** 10.1101/2025.02.25.25322862

**Authors:** Andrea Hoyos-Domingo, Fátima Ruiz-López, Belén García-Bueno, María Magdalena de la Torre-Álamo, Sandra V. Mateo, Daniel Vidal-Correoso, Pablo Luis Guzmán Martínez-Valls, Alicia López-Abad, Francisco García-Rivas, Pedro López-Cubillana, Alberto Baroja-Mazo

**Author notes:** Those authors contributed equally to this work (Andrea Hoyos-Domingo, Fátima Ruiz-López, Belén García-Bueno). Corresponding author: Alberto Baroja-Mazo. Campus de Ciencias de la Salud. Edificio LAIB. Office 4.21. Ctra. Buenavista s/n. 30120 Murcia, Spain.

## Abstract

**Background:** Kidney transplantation is the preferred treatment for end-stage renal disease, but delayed graft function remains a significant complication. Cold ischemia during organ preservation can lead to the release of danger-associated molecular patterns (DAMPs), which may influence graft outcomes. This study aimed to quantify DAMPs in kidney preservation fluid and assess their correlation with delayed graft function (DGF).

**Methods:** Preservation fluid samples from 88 deceased kidney donors were analyzed for various DAMPs, including mitochondrial DNA (mitDNA), cytochrome c, nucleosomes, hyaluronan, and inflammasome-related molecules (IL-18 and IL-1β). The influence of donor type (DBD vs. DCD) and cold ischemia time (CIT) on DAMP concentrations was evaluated. Additionally, the correlation between DAMP levels and DGF was assessed.

**Results:** Multiple DAMPs were detected in preservation fluid, including mitDNA, cytochrome c, nucleosomes, and hyaluronan. The type of donation (DBD vs. DCD) had minimal impact on DAMP concentrations, except for HSP70, which was significantly higher in DCD donors. CIT positively correlated with hyaluronan and nucleosome levels. Cytochrome c emerged as a potential biomarker for DGF, showing a significant increase in patients with early dysfunction and correlating with post-transplant creatinine levels.

**Conclusions:** Quantifying DAMPs in kidney preservation fluid is feasible and may provide valuable insights into graft quality and early post-transplant outcomes. Cytochrome c, in particular, shows promise as a biomarker for predicting delayed graft dysfunction. These findings highlight the importance of minimizing cold ischemia time and suggest that DAMP analysis could improve graft assessment prior to transplantation.

## Introduction

Kidney transplantation is the treatment of choice for patients with end-stage renal disease, offering improved quality of life and survival rates compared to dialysis [1]. However, delayed graft function (DGF) remains a significant challenge, often leading to graft loss if not promptly addressed [2]. The success of transplantation depends on multiple factors, including the quality of the donor organ and the preservation methods used.

Cold ischemia, a standard preservation technique, reduces metabolic activity but can cause endothelial and microcirculatory damage [3]. In donors after circulatory death (DCD), additional warm ischemia exacerbates organ damage due to oxygen deprivation [4]. Recent advancements in preservation technologies, such as normothermic regional perfusion (NRP) and hypothermic/normothermic machine perfusion, have improved organ viability [5, 6], but the release of danger-associated molecular patterns (DAMPs) during ischemia remains a concern.

DAMPs, including mitochondrial DAMPs, are released from damaged cells and can activate the innate immune system, leading to inflammation and graft injury [7]. The inflammasome, particularly NLRP3, plays a crucial role in this process, amplifying the inflammatory response and contributing to graft dysfunction [8].

This study aimed to quantify DAMPs in kidney preservation fluid and evaluate their correlation with donor type, cold ischemia time, and DGF. We hypothesized that DAMP levels could serve as biomarkers for graft quality and predict early post-transplant outcomes.

## Materials and Methods

### Patients and Sample Collection

Preservation fluid samples were collected from 88 deceased kidney donors at the Hospital Clínico Universitario Virgen de la Arrixaca (HCUVA, Murcia, Spain). Donors were classified as brain-dead donors (DBD) or donors after circulatory death (DCD). The study included adult patients and ran from April 2023 to April 2024, and received approval from the HCUVA (2023-1-2-HCUVA). Prior to enrollment, written consent was obtained from each participant. The study adhered to all ethical principles outlined in the Helsinki Declaration. The clinical and demographic data of both donors and recipients were collected using an electronic case report form (Table 1).

**Table 1.** Clinical and demographic data of donors and recipients included in the study.

|  | Donors (N = 88) | Recipients (N = 88) |
| --- | --- | --- |
| Age (years) | 53.58 ± 14.63; 57 (8-77) | 57.04 ± 14.03; 59 (0-80) |
| Sex |  |  |
| Male | 54 (61.36 %) | 48 (54.55 %) |
| Female | 34 (38.64 %) | 40 (45.45 %) |
| Type of donation |  |  |
| DBD | 57 (64.77 %) |  |
| DCD | 31 (35.23 %) |  |
| SRR | 0 (0 %) |  |
| NRP | 31 (100 %) |  |
| CIT (min) | 360.76 ± 248.5; 300 (90-730) |  |
| AR |  |  |
| Yes |  | 11 (12.5 %) |
| No |  | 77 (87.5 %) |
| Delayed graft function |  |  |
| Yes |  | 33 (37.5 %) |
| No |  | 55 (62.5 %) |
| Post-transplant Creatinine levels (mg/dL) |  |  |
| 1 month |  | 1.71 ± 1.07; 1.5 (0-6.79) |
| 3 months |  | 1.36 ± 0.67; 1.35 (0-3.4) |
| 6 months |  | 1.3 ± 0.67; 1.3 (0-3) |

Quantitative variables are presented as mean ± standard deviation (SD) or median (range). Qualitative variables are presented as frequency (percentage). DBD: Donor after brain death; DCD: Donor after circulatory death; SRR: Super-rapid recovery; NRP: Normothermic regional perfusion; CIT: Cold ischemia time; AR: acute rejection.

After static cold storage, donor kidneys were perfused through the renal artery with saline solution supplemented with 20 U/mL of heparin. The first 50 mL of intravascular perfusate exiting through the renal vein were collected under sterile conditions. This end-ischemic organ preservation solution (eieiOPS) was then centrifuged at 400×g for 5 minutes, aliquoted, and ultra-frozen at −80°C until further use.

### Quantification of DAMPs in eiOPS

Mitochondrial DNA (mitDNA) was extracted using the DNeasy Blood & Tissue Kit (Qiagen) and quantified by real-time PCR (RT-qPCR) using primers for the MT-CO1 gene [9]. A standard curve was generated using serial dilutions of mitDNA to quantify the samples.

Total DNA was quantified using the PicoGreen dsDNA Quantitation Kit (Invitrogen). A standard curve was generated using serial dilutions of dsDNA, and fluorescence was measured at 480 nm excitation and 520 nm emission.

Mitochondria was quantified by flow cytometry using MitoTracker Red FM and calcein-AM staining [9]. Samples were incubated at 37°C for 20 minutes, centrifuged, and resuspended in cytometry buffer. Data were acquired using a BD FACSCanto II flow cytometer and analyzed with BD FACSDiva software.

Other DAMPs (HSP70, Fibronectin, Hyaluronan, Nucleosomes, Cytochrome c or NFPs) were quantified using ELISA kits following the manufacturers’ protocols, and absorbance was measured at 450 nm using a Synergy Mx plate reader (BioTek).

### Statistical Analysis

Data were analyzed using non-parametric tests (Mann-Whitney U test) for non-normally distributed variables and Spearman’s correlation for assessing relationships between variables. Statistical significance was set at *p* ≤ 0.05. All analyses were performed using GraphPad Prism 9.0 and SPSS 23.0.

## Results

### Different DAMPs can be detected in eiOPS after cold ischemic storage of the kidney

During cold ischemia, various endogenous molecules are released into the extracellular environment. These molecules can act as DAMPs, triggering immune system activation. Therefore, we analyzed the presence of different DAMPs in eiOPS collected from donated kidneys following cold ischemic storage (Table 2). Notably, molecules associated with inflammasome signaling, such as IL-18 and IL-1β, were detected in eiOPS (Table 2). Additionally, nucleosomes, HSP70, mitochondrial DNA, total DNA, cytochrome c, fibronectin, N-formilated peptides (NFPs), hyaluronan, and free mitochondria were also identified in eiOPS (Table 2). The concentrations of these DAMPs varied widely, with mitDNA and cytochrome c showing significant variability.

**Table 2.** Quantification of various DAMPs and inflammasome-related cytokines detected in eiOPS after static cold ischemic storage.

| **DAMPs** | **Concentration** |
| --- | --- |
| Nucleosomes (RU/mL) | 1,12 (0,44-4,73) |
| HSP70 (pg/mL) | 1,32 (0-10,43) |
| Mitochondrial DNA (pg/mL) | 1,28 (0,141-7,05) |
| Total DNA (pg/mL) | 617,17 (117,7-2146,1) |
| Cytochrome c (ng/mL) | 0,183 (0,064-0,515) |
| Fibronectin (ng/mL) | 18,32 (0-258,7) |
| NFPs (pg/mL) | 263,18 ± 72,35 |
| Hyaluronan (ng/mL) | 249,83 (0-1545,97) |
| Mitochondria/mL | 1275983 (92178-5979964) |
| **Inflammasome-related cytokines** | **Concentration** |
| IL-18 (pg/mL) | 6,49 (2,67-18,18) |
| IL-1β (pg/mL) | 2,35 (0,97-5,65) |

Variables are expressed as mean ± SD for normally distributed data and as median (range) for non-normally distributed data. RU: Relative units; NFP: N-formilated peptides.

### The type of donation and CIT have a moderate impact on DAMP concentrations in eiOPS after cold ischemic storage in kidney transplantation

The type of organ donation had a minimal effect on the concentrations of DAMPs detected in eiOPS after cold ischemic storage. Notably, only HSP70 showed a significant increase in organs from DCD donors compared to those from DBD donors (Figure 1a). Likewise, no significant differences were observed for other inflammasome-related cytokines based on donor type (Figure 1b).

**Figure 1.**
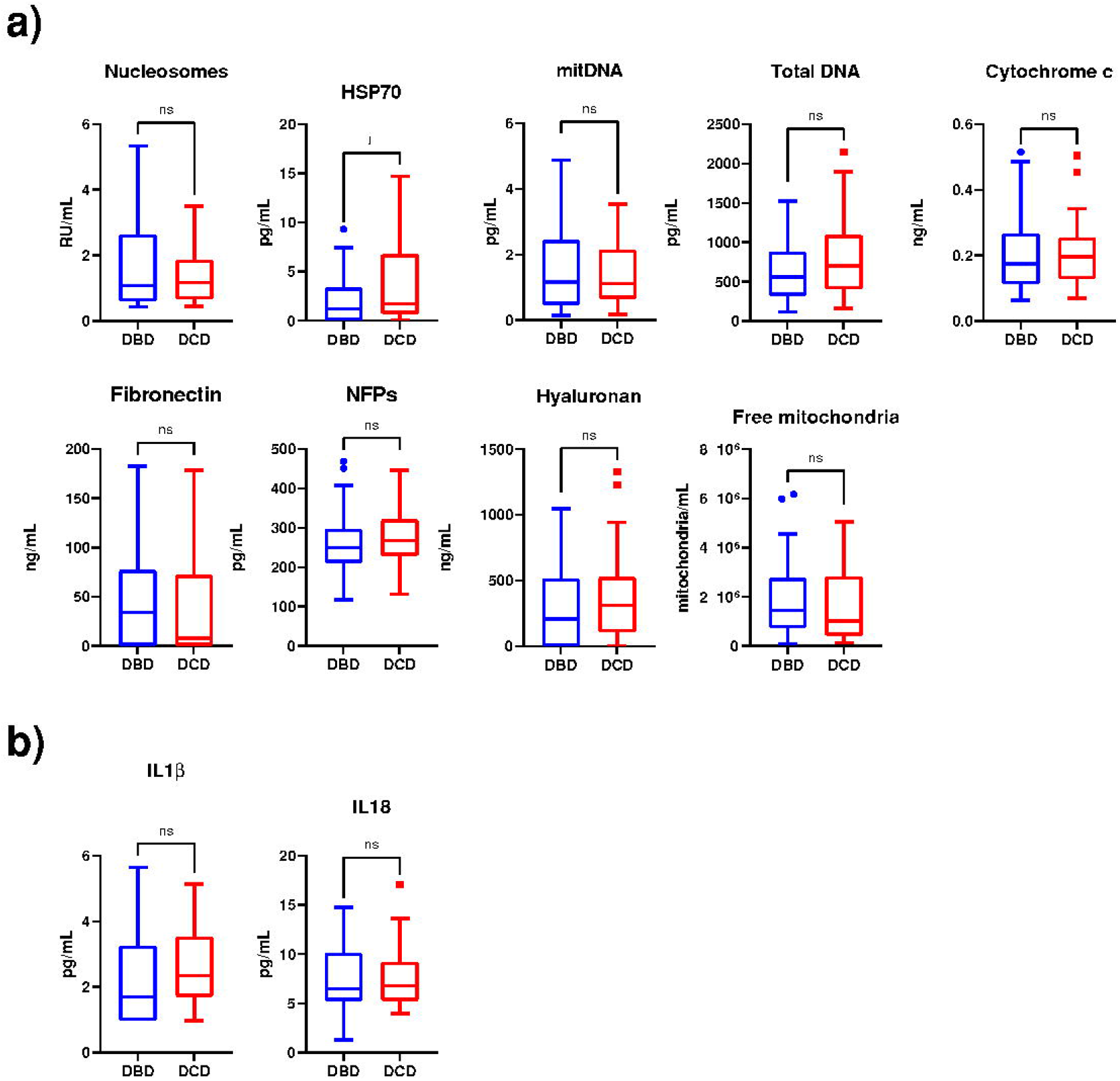
HSP70 is more highly expressed in eiOPS from DCD kidneys. Graphical representation of various DAMPs (a) and inflammasome-related cytokines (b) detected in eiOPS, categorized by donor type. Results are presented as median, interquartile range, minimum, and maximum. *p ≤ 0.05; ns = non-significant (Mann-Whitney test).

Likewise, CIT positively correlated only with hyaluronan (r = 0.24, p ≤ 0.049) and nucleosome levels (r = 0.33, p ≤ 0.005) (Figure 2a). Moreover, both type of donations presented similar CITs (Figure 2b).

**Figure 2.**
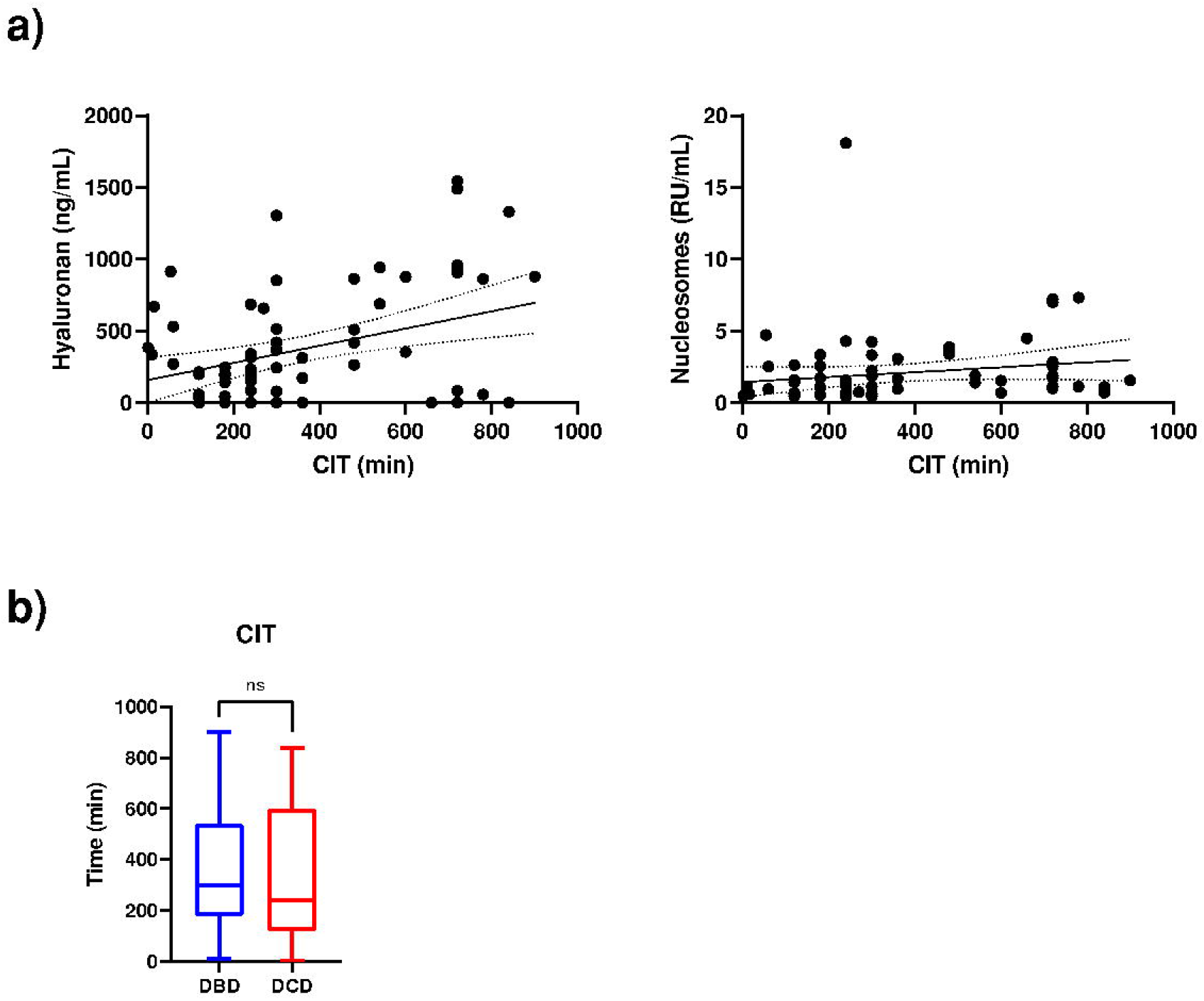
Hyaluronan and nucleosomes in eiOPS positively correlate with cold ischemia time (CIT). (a) Spearman’s correlation analysis. (b) Graphical representation of CIT, categorized by donor type. Results are presented as median, interquartile range, minimum, and maximum. ns = non-significant (Mann-Whitney test).

### Cytochrome c emerges as a potential biomarker for delayed transplant function

Among all the DAMPs analyzed, only cytochrome c was significantly elevated in patients with delayed function (Figure 3a). Furthermore, its correlation with creatinine levels, a key marker for assessing renal function post-transplant, was evaluated at one, three, and six months after transplantation, revealing a positive correlation (r = 0.33, p = 0.002 at 1 month; r = 0.246, p = 0.022 at 3 months; r = 0.212, p = 0.048 at 6 months) (Figure 3b).

**Figure 3.**
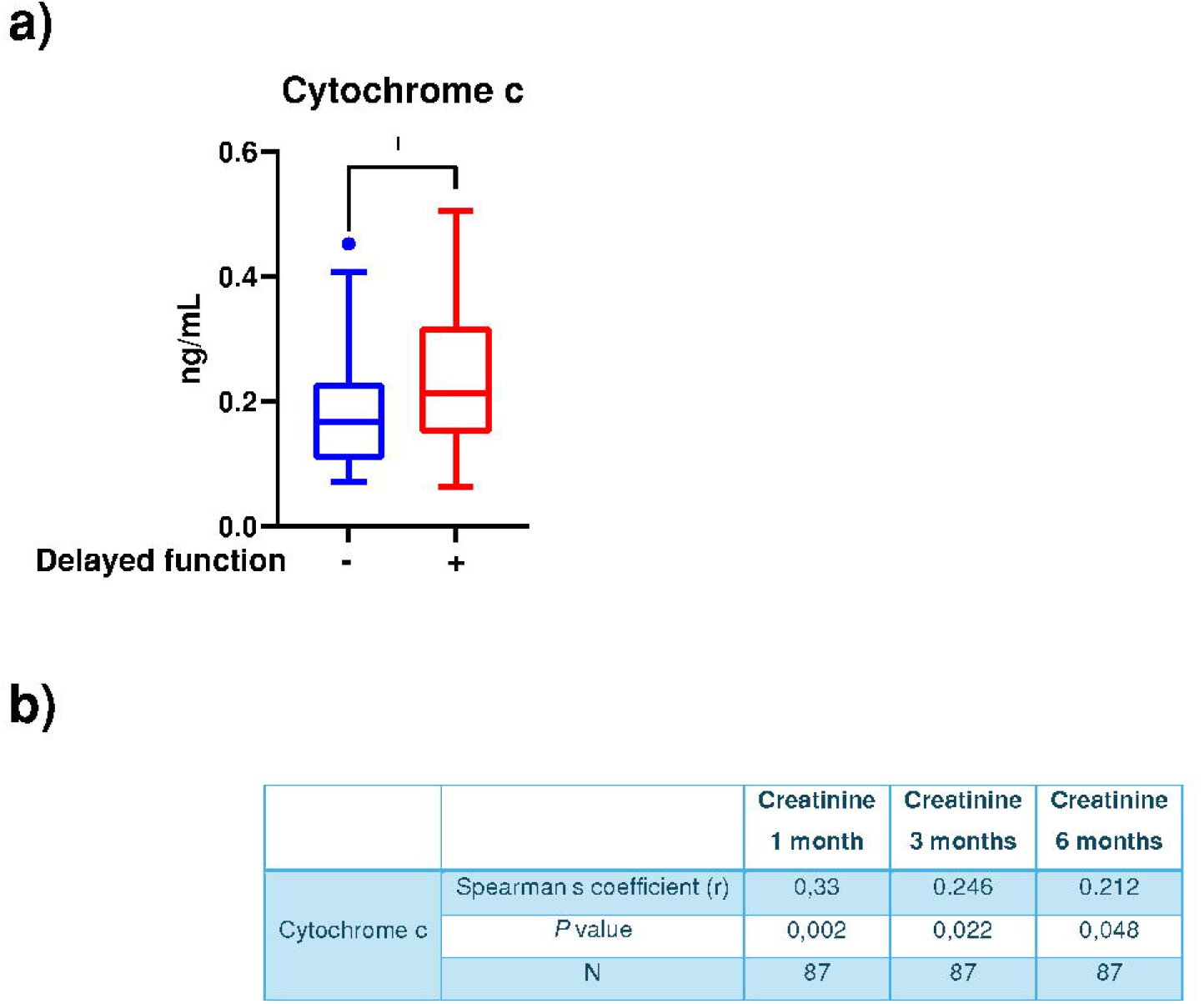
Cytochrome c levels in eiOPS correlate with post-transplant creatinine and delayed graft function (DGF). (a) Graphical representation of cytochrome c levels in patients with and without DGF. Results are presented as median, interquartile range, minimum, and maximum. *p ≤ 0.05 (Mann-Whitney test). (b) Correlation matrix between cytochrome c concentration in eiOPS and recipient blood creatinine levels at three different time points post-transplantation.

## Discussion

This study highlights the feasibility of quantifying DAMPs in kidney preservation fluid and their potential role in predicting DGF. The detection of DAMPs such as mitDNA, cytochrome c, and nucleosomes underscores the impact of cold ischemia on organ quality.

Disparities related to donor type appear to influence not only this specific organ but also the broader context of organ transplantation as a whole [10]. Studies on donation type suggest that in DCD, the warm ischemia period correlates with a higher number of necrotic cells during reperfusion, leading to increased DAMP release and a heightened immune response, as previously reported by our group in liver transplantation [7]. However, in the present study, no significant differences were observed in the analyzed variables concerning donor type. A key difference, beyond the organ studied, is that in the former, only 26% of DCD organs underwent normothermic regional perfusion (NRP) [7], whereas in the current study, 100% of these organs were obtained using this technique. Traditionally, DCD organs posed greater challenges due to the warm ischemia period before procurement, potentially affecting their viability. However, with advancements in preservation techniques such as NRP and hypothermic or normothermic machine perfusion (HMP/NMP), these differences have been significantly reduced [11].

Many authors have reported that longer ischemia durations are associated with an increased risk of early dysfunction and acute rejection [12, 13]. In our study, significant correlations were observed between cold ischemia time and hyaluronan and nucleosome levels, indicating greater kidney damage with prolonged ischemia. Particularly, cold ischemia time has been identified as an independent predictor of graft loss in DCD organs obtained through NRP [14]. For instance, in kidney transplantation, prolonged cold ischemia has been linked to suboptimal renal function and reduced graft survival, regardless of donor type [15].

Notably, cytochrome c emerged as a promising biomarker for DGF, with elevated levels correlating with poorer renal function post-transplantation. Cytochrome c, a mitochondrial protein released during cellular stress, has been implicated in the activation of the innate immune system and the amplification of inflammatory responses [16]. In this study, we observed significantly higher levels of cytochrome c in patients with DGF, suggesting that its release during cold ischemia may contribute to post-transplant complications. The positive correlation between cytochrome c levels and post-transplant creatinine levels further supports its potential as a predictive biomarker for graft outcomes.

The role of cytochrome c in graft dysfunction may be mediated through its ability to activate endothelial cells and promote the release of pro-inflammatory cytokines, such as IL-6 and TNF-α [17]. Additionally, cytochrome c can induce the production of reactive oxygen species (ROS), which exacerbate tissue damage and inflammation [18]. These mechanisms align with previous studies showing that mitDAMPs play a critical role in the inflammatory response following ischemia-reperfusion injury [19, 20].

The quantification of cytochrome c in preservation fluid could provide clinicians with a valuable tool for assessing graft quality prior to transplantation. By identifying grafts with elevated cytochrome c levels, transplant teams could implement targeted interventions, such as antioxidant therapies or anti-inflammatory treatments, to mitigate the risk of early dysfunction. Furthermore, the correlation between cytochrome c and post-transplant creatinine levels suggests that this biomarker could be used to monitor graft function in the early post-transplant period.

Future research should include a larger cohort and explore the role of other DAMPs and inflammatory pathways. Additionally, the development of standardized protocols for DAMP quantification in preservation fluid will be essential for the clinical implementation of these findings.

## Data Availability

Ethics committee/IRB of Full Institution name gave ethical approval for this work

## Notes

### Competing Interest Statement

The authors have declared no competing interest.

### Funding Statement

This study was funded by Instituto de Salud Carlos III (DTS23/00013) and Fundacion Seneca de la Region de Murcia (22257/PDC/23)

### Author Declarations

Ethics committee of Hospital Clinico Universitario Virgen de la Arrixaca gave ethical approval for this work (2023-1-2-HCUVA)

